# Effectiveness and safety of reactive focal mass drug administration (rfMDA) using dihydroartemisinin-piperaquine to reduce malaria transmission in very low-endemic setting of Eswatini: a pragmatic cluster randomised controlled trial

**DOI:** 10.1101/2021.03.12.21252721

**Authors:** Sibonakaliso Vilakati, Nontokozo Mngadi, Jade Benjamin-Chung, Nomcebo Dlamini, Mi-Suk Kang Dufour, Brooke Whittemore, Khayelihle Bhangu, Lisa M. Prach, Kimberly Baltzell, Nomcebo Nhlabathi, Calisile Malambe, Bongani Dlamini, Danica Helb, Bryan Greenhouse, Gugu Maphalala, Deepa Pindolia, Muhindo Kalungero, Getahun Tesfa, Roly Gosling, Nyasatu Ntshalintshali, Simon Kunene, Michelle S. Hsiang

**Affiliations:** Eswatini National Malaria Program, Manzini, Eswatini; Clinton Health Access Initiative, Mbabane, Eswatini; Division of Epidemiology & Biostatistics, University of California, Berkeley, USA; Malaria Elimination Initiative, Global Health Group, UCSF; Department of Medicine, University of California, San Francisco (UCSF), USA; Department of Pediatrics, University of Texas Southwestern Medical Center, Dallas, USA; Department of Family Health Care Nursing, UCSF; Department of Medicine National Clinical Laboratory Services, Mbabane, Eswatini; Good Shepherd Hospital, Siteki, Eswatini; Raleigh Fitkin Memorial Hospital, Manzini, Eswatini; Department of Pediatrics, UCSF

**Keywords:** Plasmodium, Swaziland, low transmission, malaria elimination, active case detection, reactive case detection, antimalarial, dihydroartemisin-piperaquine, cluster randomized controlled trial

## Abstract

**Introduction:** To reduce malaria transmission in very low-endemic settings, screening and treatment near index cases (reactive case detection (RACD)), is widely practiced, but the rapid diagnostic tests (RDTs) used miss low-density infections. Presumptive treatment near index cases (reactive focal mass drug administration (rfMDA)) may be safe and more effective.

**Methods:** We conducted a cluster-randomised controlled trial in Eswatini, a very low-endemic setting. 77 clusters were randomised to rfMDA using dihydroartemisin-piperaquine (DP) or RACD involving RDTs and artemether lumefantrine (AL). Interventions were delivered by the local programme. An intention-to-treat analysis was used to compare cluster-level cumulative confirmed malaria incidence among clusters with cases. Secondary outcomes included safety and adherence.

**Results:** From Sept 2015–Aug 2017, 220 index cases from 47 clusters triggered 49 RACD events and 68 rfMDA events. RACD and rfMDA were delivered to 1696 and 1932 individuals, respectively. Index case and target population intervention coverages for both arms were 75.6%–81.4% and adherence to DP was 98.7%. For rfMDA versus RACD, cumulative incidences (per 1000 person-years) of all malaria were 2.11 (95% CI 1.73–2.59) and 1.97 (1.57–2.47), respectively; and of locally acquired malaria, they were 1.29 (95% CI 1.00–1.67) and 0.97 (0.71–1.34), respectively. Adjusting for imbalance in baseline incidence, incidence rate ratio (aIRR) for rfMDA versus RACD was 0.93 (95% CI 0.54–1.60) for all malaria and 0.77 (95% CI 0.38–1.56) for locally acquired malaria. No serious adverse events occurred.

**Conclusion:** In a very low-endemic, real-world setting, this trial is the first to evaluate rfMDA using DP. rfMDA was safe and resulted in lower cumulative incidence compared to RACD, but we were unable to confirm its effectiveness, potentially due to insufficient power. To assess impact of interventions in very low-endemic settings, multi-site, adaptive trials and use of complementary interventions may be needed.

**What is already known?:**

- Reactive case detection (RACD), or malaria testing and treatment in the vicinity of passively detected malaria cases, is a standard of care intervention used in low and very low transmission settings aiming for malaria elimination.
- Despite the use of RACD, progress toward malaria elimination has stalled in many countries and new strategies are needed.
- Reactive focal mass drug administration (rfMDA) is a transmission reducing strategy that has been shown to be effective in a low transmission setting, but there are no trial data from a very low transmission setting.

**What are the new findings?:**

- In a pragmatic, cluster-randomised controlled trial of rfMDA using dihydroartemisinin-piperaquine compared to RACD, we found that rfMDA was safe.
- rfMDA resulted in lower cumulative incidence, but we were unable to confirm its effectiveness compared to RACD, potentially due to insufficient power (we expected 63 total clusters would have incident cases, but observed 47).

**What do the new findings imply?:**

- When implemented in a real-world, very low transmission setting, rMDA was safe but evidence regarding its effectiveness to reduce transmission was weak.
- The challenge to show a statistically significant impact of a targeted community-based intervention in a very low transmission setting highlights the need for such trials to be multi-site, adaptive, and consider use of complementary interventions.

## Background

Since 2000, many countries have scaled up effective malaria control interventions, resulting in reductions in malaria burden and a renewed goal to eradicate malaria worldwide by 2050.(1) When the goal is to interrupt transmission, it may be necessary to treat not only symptomatic malaria but also asymptomatic infections which perpetuate ongoing transmission and represent an increasing proportion of all infections in low transmission settings.(2, 3)

To address asymptomatic infections, one widely practiced strategy is active case detection in household members and neighbours of symptomatic cases recently reported from health facilities, also known as reactive case detection (RACD).(4) Since malaria infections cluster in space and time,(3) RACD can target limited resources to areas at highest risk of infection. In settings with substantial imported malaria cases that may seed local transmission, RACD also serves as a focal outbreak response.(4) However, the effectiveness of RACD is limited by the low sensitivity of currently available point-of-care diagnostics to detect low-density and non-falciparum infections. Molecular testing such as polymerase chain reaction (PCR) or loop-mediated isothermal amplification (LAMP) improves sensitivity but is not practical given costs, logistical challenges of specimen collection and transport, and turn-around time required for laboratory testing and return visits to treat test-positive individuals. In addition, mass screening and treatment, which is similar to RACD but delivered community-wide, has not sustainably reduced incidence in prior studies.(5) As such, the World Health Organization (WHO) does not recommend RACD as a strategy to reduce or interrupt transmission.

Mass drug administration (MDA), or the treatment all individuals within a specified area with an effective antimalarial irrespective of infection status,(6, 7) may address some of the challenges of RACD. MDA was a component of many malaria elimination programmes in the mid-twentieth century but fell out of favor due to concerns regarding its effectiveness, sustainability, cost, and fear of accelerating drug resistance. More recent evidence suggests that when implemented in areas of low endemicity and in combination with other interventions, MDA has the potential to sustainably interrupt transmission.(6, 7) Maximizing coverage and adherence may also help to mitigate risks of drug resistance.(8) MDA has recently been recommended by the WHO in areas approaching interruption of transmission where there is good access to treatment, effective implementation of vector control and surveillance, and minimal risk of re-introduction of infection.(9) However, a dearth of definitive evidence on its effectiveness, safety, and feasibility remains.(10)

Eswatini (formerly Swaziland) is among 21 countries worldwide that were identified by WHO as the most likely to reach zero indigenous cases by 2020.(11) However, several of these countries including Eswatini continue to experience persistent local transmission and resurgence. As a malaria elimination-specific strategy, the Eswatini National Malaria Programme (NMP) has implemented RACD since 2009. Prior studies have confirmed that asymptomatic infections cluster around passively detected index cases, with the highest risk within 200 meters of the index case.(12) However, in Eswatini RACD using RDTs missed two-thirds of infections and 40% of hotspots compared to more sensitive molecular methods.(12) Due to logistical challenges, attempts to use more sensitive molecular methods to directly inform treatment have been unsuccessful (N. Dlamini, personal communication).

Reactive focal MDA (rfMDA) is an alternative intervention that builds on RACD for targeting high-risk populations residing near index cases. rfMDA entails mass drug administration without testing in household members and neighbours of recent index cases.(13) A recent trial of rfMDA using artemether-lumefantrine (AL) from a low transmission setting (infection prevalence 1–10%(14)) with minimal importation in Namibia reported safe administration and rfMDA reduced locally acquired malaria incidence by approximately 50% compared to RACD.(13) However, there are no trials of rfMDA from very low transmission settings (infection prevalence >0 but <1%(14)) with a high level of importation, which characterizes most near-elimination settings. rfMDA may be more appropriate than blanket MDA in low-endemic settings, since it targets populations where malaria has been recently introduced. Also, there are no trials of rfMDA using dihydroartemisinin-piperaquine (DP), which compared to standard artemisinin-combination therapies such as AL, has favorable characteristics for MDA (less frequent dosing and longer period of protection), but safety concerns about rare QT-interval prolongation leading to arrythmia and sudden death exist.(15)

Our objective in this trial was to evaluate the effectiveness of rfMDA using DP, compared to RACD, for reducing malaria transmission in the very low transmission setting of Eswatini. Both the rfMDA and RACD interventions were embedded within the Eswatini National Malaria Programme; as such, this pragmatic trial assessed real-world effectiveness of these interventions when delivered within an existing surveillance and response programme.

## Methods

### Study design and participants

We conducted a pragmatic open-label, cluster-randomised controlled trial(16) between September 2015 and June 2017 in the Kingdom of Eswatini, a low middle-income country in southern Africa. Approximately 30% of the population lives in the eastern malaria-endemic area, which borders Mozambique. *Plasmodium falciparum* is responsible for over 99% of malaria cases in Eswatini. Malaria transmission is unstable and occurs mainly between October and May.(12) Annual case loads are reported from July to June each year.

After major declines in malaria transmission from annual parasite incidence (API) of 3.9 to 0.07 per 1000 population from 1999 to 2009, the NMP reoriented its strategy from control to elimination of transmission by 2020. Since implementation of the elimination programme and until just prior to this trial, API has remained <1 per 1000 population. In 2014–2015, the transmission season prior to the trial, there were 604 reported cases, of which 50% were classified as imported.(17)

This pragmatic cluster-randomised controlled trial was designed to compare rfMDA and RACD effectiveness as implemented by the Eswatini National Malaria Programme (NMP) and in the context of other ongoing interventions including case management, vector control, surveillance, and information, communication and education. 77 malaria endemic localities or clusters, with a total of 209085 individuals residing in 431 enumeration areas, were eligible for inclusion. Of the 77 clusters, 63 had malaria cases in the three years prior to the trial; the remainder did not have cases but had prior historical risk of malaria transmission. We randomised clusters with a 1:1 allocation ratio to receive RACD, including rapid diagnostic testing with *Pf-*specific First Response (Premier Medical Corporation Ltd, Mumbai, India) and treatment of positives with AL (Coartem, Novartis Pharmaceuticals, Kempton Park, South Africa) or rfMDA with presumptive treatment using DP (Eurartesim, Sigma Tau, Italy) (Figure 1). Inclusion and exclusion criteria are shown in Appendix 1. Briefly, microscopy- or RDT-confirmed index cases reported from any health facility in Eswatini were classified as local, imported, or unknown based on travel history. RACD or rfMDA was triggered if the index case resided within a study cluster. If RACD was conducted in the prior 5 weeks of the index case report, it was not repeated. Following the manufacturer’s recommendation that DP not be repeated within 8 weeks, nor taken more than twice in a year, rfMDA was not repeated if these criteria were met. Other exclusion criteria for DP included: age < 9 months; weight < 7 kg; pregnancy and breastfeeding, allergy to DP, acute illness including severe malaria, underlying kidney or hepatic problems, personal or family history of QT prolongation, or recent treatment with QT-prolongating medications.

**Figure 1.**
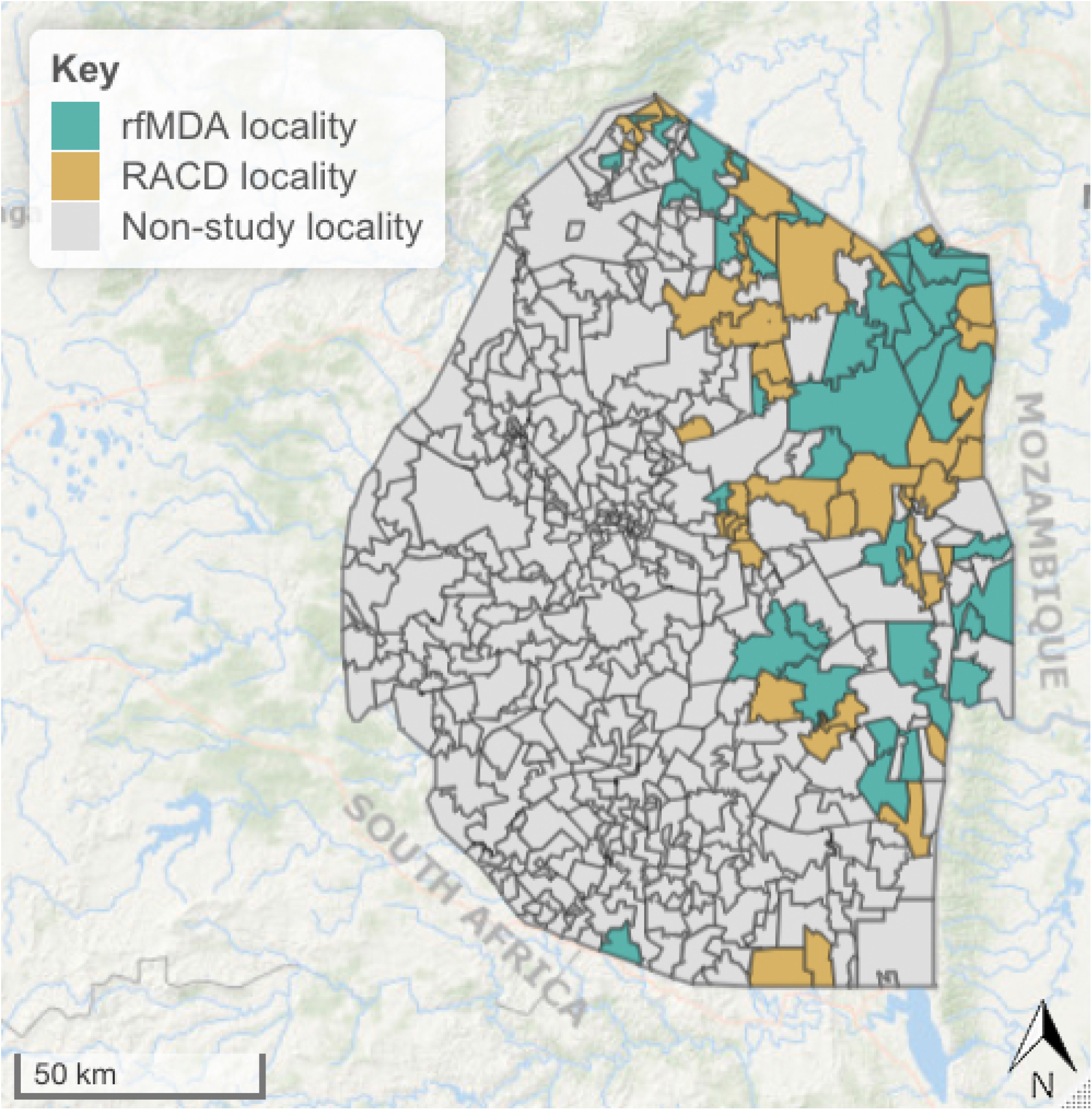
Map of the study area Abbreviations: rfMDA, reactive focal mass drug administration; RACD, reactive case detection.

### Randomisation and masking

To ensure that the baseline risk of malaria was balanced between intervention arms, we utilized block-stratified randomisation. We assigned the 77 localities or clusters to randomisation blocks by separating them into three risk groups based on incidence in the three years prior to the trial and prior historical risk according to NMP. We further stratified each block by whether the size of the population at risk was above or below 650 individuals. A statistician at UC San Francisco (MKD) generated the random allocation sequence using SAS (version 9.4m2) to randomly assign 0 or 1 to each cluster within each block and stratum, and the NMP flipped a coin to determine which intervention corresponded to 0 and 1. The intervention delivery team and study investigators were not blinded to intervention assignment due to the nature of interventions.

### Procedures

Prior to the study, individuals residing in endemic areas received indoor residual spraying (IRS) per standard procedures by the NMP. During the study, malaria cases who presented at surveillance sites were confirmed using RDT or microscopy. Index cases triggered interventions if they lived within the study area. The surveillance team attempted to visit index cases’ homes within 48 hours to administer a questionnaire about travel history and vector control measures.

In the RACD arm, consistent with NMP standard practices, all consenting individuals residing within 500m of the index case (the “target population” for RACD) received RDT testing, and a dried blood spot (DBS) was collected for subsequent molecular testing. RDT-positive individuals were transported to the nearest health facility for treatment. The study aimed to deliver interventions within 7 days of index case presentation, but allowed up to 5 weeks.

In the rfMDA arm, individuals residing within 200m of the index case, but extending beyond 200m to reach a minimum of 30 individuals (the “target population” for rfMDA) were targeted for drug administration using DP. A radius of 200 meters with a minimum 30 individuals was chosen because prior RACD studies showed that the majority of infections near an index case could be captured within this target population.(12) Field staff assessed whether it was safe to administer DP to enrolled eligible individuals. Individuals ineligible to receive DP received RDT testing, and a DBS was collected for subsequent molecular testing. RDT-positive individuals were transported to the nearest health facility for treatment. Eligible individuals received the first dose of DP under directly observed therapy and doses for day 2 and day 3 for self-administration. Individuals ineligible for DP were screened using RDTs and transported to the nearest health facility for treatment if they tested positive. Participants were instructed to go to the nearest health facility if they experienced any illness after taking DP, and they were instructed to contact an on-call study nurse that was available at all hours. To assess adherence, the study team returned to a subsample of participants (all participants of the first intervention for each rfMDA cluster) 7-10 days after enrollment to conduct pill counts.

In both arms, study teams returned a second and third day to recruit individuals who were initially absent. The study aimed to achieve at least 80% intervention coverage of index cases and 80% coverage of the target population.

### Laboratory methods

RDT testing was performed using the First Response *P. falciparum* HRP-2 Detection Test (Premier Medical Corporation Ltd.). DNA extraction from DBS for LAMP testing was conducted as previously described (Loopamp Malaria Pan and Pf Detection Kits, Eiken Chemical Co., Ltd.).(12) LAMP results were used for research purposes only.

### Outcomes

The primary outcome of the trial was the cumulative incidence of malaria cases by study cluster over two-years of follow-up. Secondary outcomes reported here include safety and adherence (acceptability has been reported elsewhere(18)). Infection prevalence and seroprevalence at two-year follow-up were originally also secondary outcomes but the endline cross-sectional survey was not conducted due to a shift in priorities within the Ministry of Health.

### Statistical Analysis

We estimated the minimum detectable difference in cumulative incidence per population at-risk between arms. Based on surveillance data from 2012–2015 in areas where RACD was conducted, we assumed an annual incidence of 4 per 1000 individuals, coefficient of variation of 0.9, and type I error of 0.05. We assumed the population at-risk (the total population of census enumeration areas that reported incident cases within each cluster) was 55 928 individuals in at least 63 of the 77 total cluster. There was 80% statistical power to detect a 50% percent reduction between arms if at least 63 of 77 clusters had at least one index case.(19)

The cumulative incidence in each cluster was calculated as the number of passively detected malaria cases divided by the product of population and follow-up time in each cluster, starting on the date of first index case detection. The first index case in each cluster was excluded from incidence calculations since interventions were delivered after initial index case detection in each cluster. Malaria-free survival was compared, and the assumption of proportional hazards was assessed using Schoenfeld residuals testing.(20)

To estimate intervention effects, we used an intention-to-treat (ITT) approach that excluded localities with no incident cases during the study period since these localities did not receive interventions. The primary analysis used negative binomial regression models with an offset for population size to estimate incidence rate ratios in each cluster over the study period. Models adjusted for baseline covariates that were associated with the outcome using a likelihood ratio test (p-value < 0.2) and that had a Pearson correlation coefficient with the outcome ≥ 0.3.(21) Baselines covariates included: incidence (2014–2015), proportion of imported cases, proportion of houses receiving IRS in the past year, monthly average enhanced vegetation index, monthly average rainfall, monthly average land surface temperature, and elevation.

Malaria transmission is highly heterogeneous in lower transmission settings(22), and trends in monthly incidence differed between arms in the three years prior to the trial. To account for these pre-trial differences, we used a synthetic control analysis to minimize pre-trial differences in incidence between arms (Appendix 2).(23) We then estimated the difference-in-differences for RACD vs rfMDA and the synthetic RACD vs rfMDA. This analysis was not pre-specified.

To assess potential contamination due to a lack of buffer zones between clusters, we identified all clusters with contiguous neighbouring clusters and plotted the incidence in each cluster against incidence in the neighbouring cluster. The small number of contiguous clusters precluded the use of formal statistical testing to assess correlations between incidences in contiguous clusters.

## Results

Between September 2015 and June 2017, 22 of the 38 clusters randomly assigned to RACD had 99 reported cases; 56 of these cases were covered by forty-nine RACD events. Twenty-five of the 39 clusters randomly assigned to rfMDA had 121 reported cases; 89 of these cases were covered by 68 rfMDA events. The remaining cases did not receive reactive interventions due to staff limitations, fuel shortages, or weather conditions complicating transport (Figure 2). Of the 2134 individuals eligible to receive RACD, 1696 (79%) were tested by RDTs. Five RDT-positive cases, of which three were LAMP positive), were referred for treatment with AL. The most common reason for non-receipt of RACD was not present (n=398, 18.7%); only 1.5% (n=33) refused. Of the 2623 individuals eligible to receive rfMDA, 1932 (74%) received DP. The most common reasons for non-receipt of rfMDA were not present (n=302, 11.5%) and ineligibility of receive DP (n=313, 11.9%) mainly due to reported potential for medication interaction. Seventy-six (2.9%) of eligible individuals refused to participate. Data on medication type were incomplete as nurses reported sensitivities around participants disclosing use of antiretrovirals (ARVs). No RDT nor LAMP-positive individuals were identified among rfMDA ineligibles. In total, 117 intervention events were implemented in the trial and 3941 individuals in 47 clusters where interventions were conducted were included in primary outcome analyses. Adherence to intervention assignment was incomplete: 20 RACD interventions were delivered in rfMDA areas (14 clusters), and 5 rfMDA interventions were delivered in RACD areas (four clusters).

**Figure 2.**
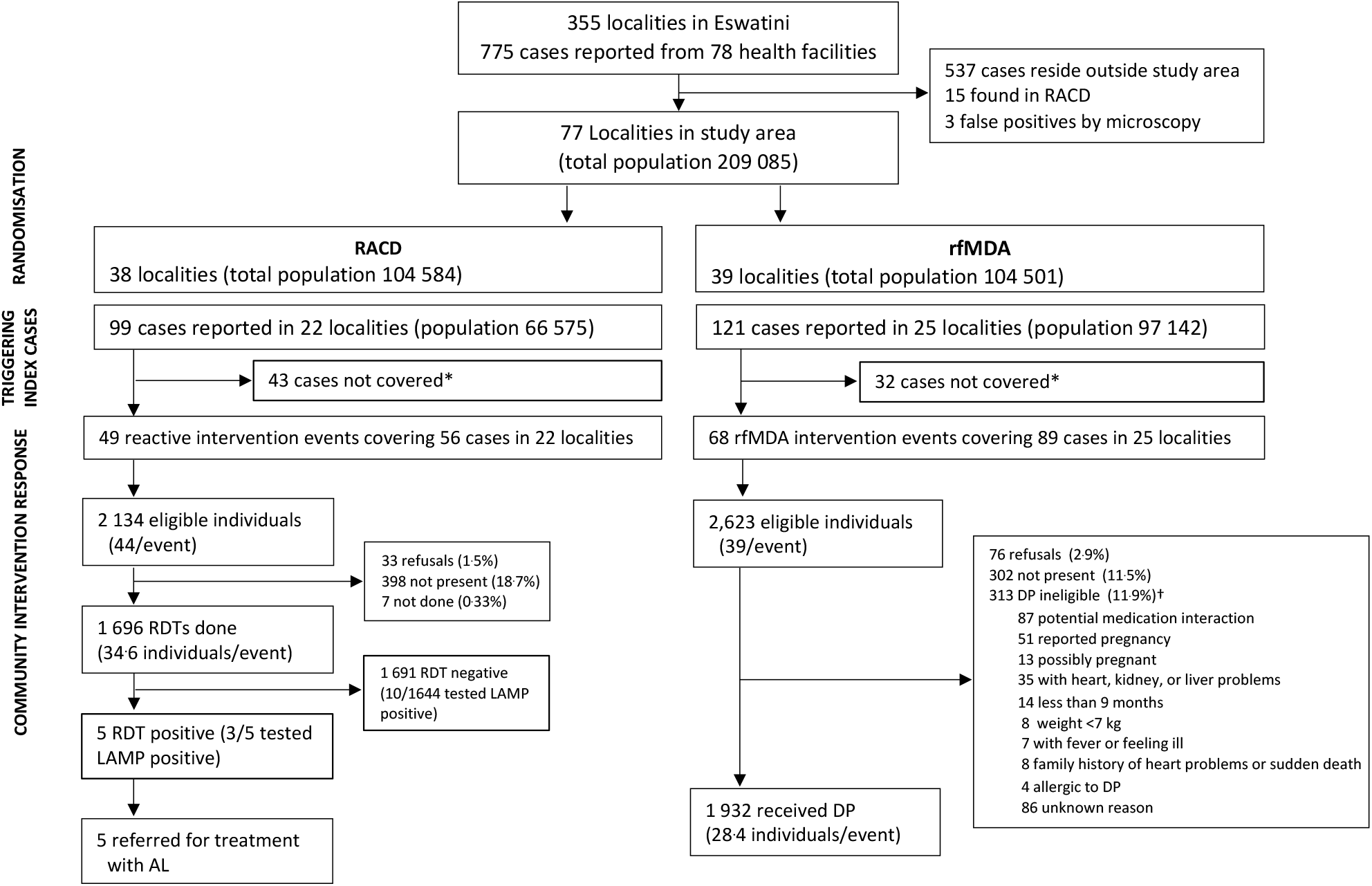
Trial profile showing randomisation and enrolment Abbreviations: rfMDA, reactive focal mass drug administration; RACD, reactive case detection; RDT, rapid diagnostic test; LAMP, loop-mediated isothermal amplification; AL, artemether-lumefantrine; DP, dihydroartemisinin-piperaquine *not covered due to staff limitations, fuel shortages, or weather conditions complicating transport †RDT testing conducted in 262 of DP ineligibles. As none tested positive, none were referred for treatment with AL

Not taking into account clustering, index cases had a similar distribution of age, sex, case origin (e.g. local, imported, or unknown), occupation, and bed net ownership between study arms (Appendix 3). The percentage of index cases that reported having had their home sprayed in the past year was higher in rfMDA clusters than RACD clusters (28.6% vs 5.3%). For target population receiving study interventions, there was a similar distribution of age, occupation, and vector control coverage. A higher proportion in the rfMDA arm (1.4%) worked in manufacturing compared to RACD (0.1%). In all study clusters, an average of 35.7% of index cases and 2.8% of the target population reported international travel in the prior 8 weeks during the study period.

Taking into account clustering, there was imbalance in baseline transmission intensity. Cumulative incidence of all malaria in the three years preceding the trial was higher in the rfMDA arm compared to the RACD arm (6.30 vs 4.17 per 1000, respectively) with a similar trend seen for local cases only, and for all and local cases only in 2014–2015, the year preceding the trial (Table 1, Appendix 4a). The percentage of cases classified as imported in each cluster in the years prior to the trial was higher in the RACD arm compared to the rfMDA arm (35.8% vs 28.1% for 2012–2015, and 48.7% vs 32.2% for 2014–2015). Mean population size and ecological factors including rainfall, enhanced vegetative index, elevation, and daytime land surface temperature were balanced between arms at baseline (Table 1).

**Table 1.**
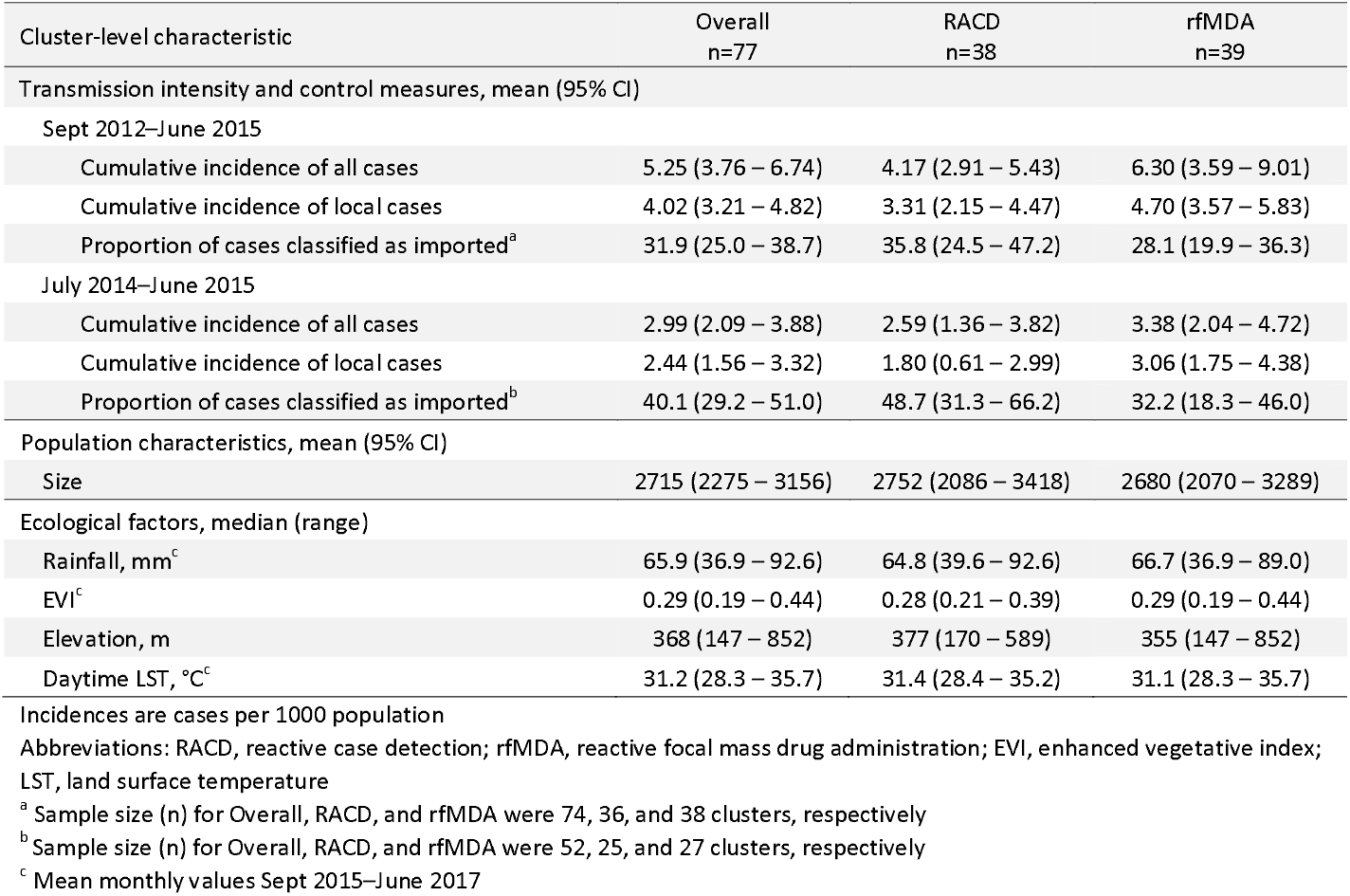
Baseline characteristics of clusters (localities) included in the trial

| Cluster-level characteristic | Overall<br>n=77 | RACD<br>n=38 | rfMDA<br>n=39 |
| --- | --- | --- | --- |
| Transmission intensity and control measures, mean (95% CI) |  |  |  |
| Sept 2012–June 2015 |  |  |  |
| Cumulative incidence of all cases | 5.25 (3.76 – 6.74) | 4.17 (2.91 – 5.43) | 6.30 (3.59 – 9.01) |
| Cumulative incidence of local cases | 4.02 (3.21 – 4.82) | 3.31 (2.15 – 4.47) | 4.70 (3.57 – 5.83) |
| Proportion of cases classified as imported<sup>a</sup> | 31.9 (25.0 – 38.7) | 35.8 (24.5 – 47.2) | 28.1 (19.9 – 36.3) |
| July 2014–June 2015 |  |  |  |
| Cumulative incidence of all cases | 2.99 (2.09 – 3.88) | 2.59 (1.36 – 3.82) | 3.38 (2.04 – 4.72) |
| Cumulative incidence of local cases | 2.44 (1.56 – 3.32) | 1.80 (0.61 – 2.99) | 3.06 (1.75 – 4.38) |
| Proportion of cases classified as imported<sup>b</sup> | 40.1 (29.2 – 51.0) | 48.7 (31.3 – 66.2) | 32.2 (18.3 – 46.0) |
| Population characteristics, mean (95% CI) |  |  |  |
| Size | 2715 (2275 – 3156) | 2752 (2086 – 3418) | 2680 (2070 – 3289) |
| Ecological factors, median (range) |  |  |  |
| Rainfall, mm<sup>c</sup> | 65.9 (36.9 – 92.6) | 64.8 (39.6 – 92.6) | 66.7 (36.9 – 89.0) |
| EVI<sup>c</sup> | 0.29 (0.19 – 0.44) | 0.28 (0.21 – 0.39) | 0.29 (0.19 – 0.44) |
| Elevation, m | 368 (147 – 852) | 377 (170 – 589) | 355 (147 – 852) |
| Daytime LST, °C<sup>c</sup> | 31.2 (28.3 – 35.7) | 31.4 (28.4 – 35.2) | 31.1 (28.3 – 35.7) |

Abbreviations: RACD, reactive case detection; rfMDA, reactive focal mass drug administration; EVI, enhanced vegetative index; LST, land surface temperature
<sup>a</sup> Sample size (n) for Overall, RACD, and rfMDA were 74, 36, and 38 clusters, respectively
<sup>b</sup> Sample size (n) for Overall, RACD, and rfMDA were 52, 25, and 27 clusters, respectively
<sup>c</sup> Mean monthly values Sept 2015–June 2017

Index case and target population intervention coverage in the RACD arm was 80.1% and 75.6%, respectively, compared to 77.0% and 81.4%, respectively, in the rfMDA arm (Table 2). Total coverage (including both index cases and the target population around each index case) was 60.0% in RACD and 68.8% in rfMDA. For all coverage measures, 95% confidence intervals for each arm overlapped substantially. The median number of days between index case report and intervention response was 7 (range: 2, 27) in the RACD arm and 11 (range: 3, 52) in the rfMDA arm. In the rfMDA arm, two clusters had response times of 40 and 52 days; excluding those clusters, the range was 3 to 21.

**Table 2.**
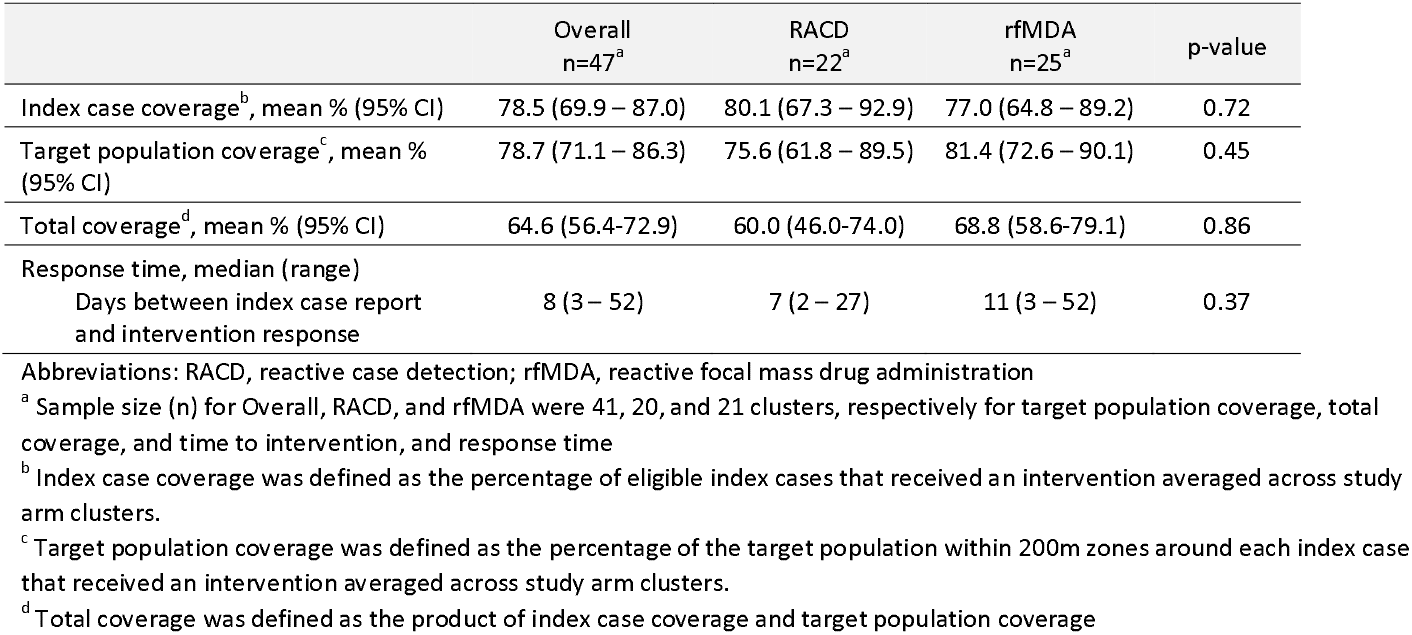
Intervention coverage and response time

|  | Overall<br>n=47<sup>a</sup> | RACD<br>n=22<sup>a</sup> | rfMDA<br>n=25<sup>a</sup> | p-value |
| --- | --- | --- | --- | --- |
| Index case coverage<sup>b</sup>, mean % (95% CI) | 78.5 (69.9 – 87.0) | 80.1 (67.3 – 92.9) | 77.0 (64.8 – 89.2) | 0.72 |
| Target population coverage<sup>c</sup>, mean % (95% CI) | 78.7 (71.1 – 86.3) | 75.6 (61.8 – 89.5) | 81.4 (72.6 – 90.1) | 0.45 |
| Total coverage<sup>d</sup>, mean % (95% CI) | 64.6 (56.4-72.9) | 60.0 (46.0-74.0) | 68.8 (58.6-79.1) | 0.86 |
| Response time, median (range) |  |  |  |  |
| Days between index case report and intervention response | 8 (3 – 52) | 7 (2 – 27) | 11 (3 – 52) | 0.37 |

Abbreviations: RACD, reactive case detection; rfMDA, reactive focal mass drug administration
<sup>a</sup> Sample size (n) for Overall, RACD, and rfMDA were 41, 20, and 21 clusters, respectively for target population coverage, total coverage, and time to intervention, and response time
<sup>b</sup> Index case coverage was defined as the percentage of eligible index cases that received an intervention averaged across study arm clusters.
<sup>c</sup> Target population coverage was defined as the percentage of the target population within 200m zones around each index case that received an intervention averaged across study arm clusters.
<sup>d</sup> Total coverage was defined as the product of index case coverage and target population coverage

During follow-up, the cumulative incidence decreased from baseline levels in both the RACD and rfMDA arms, with rfMDA having fewer cases during the final months (January to May) of the second transmission season (Figure 3, Appendix 4b). The cumulative incidence from 2015–2017 was 2.11 per 1000 in the rfMDA arm compared to 1.97 in the RACD arm (Table 3) (N = 47 clusters in both arms). In the intention-to-treat analysis, crude and adjusted incidence rate ratios (IRRs) were 1.01 (95% CI 0.58, 1.73) and 0.93 (95% CI 0.54, 1.60), respectively (Table 3). Restricting to local cases only, the adjusted IRR was 0.77 (95% CI 0.38, 1.56).

**Table 3.**
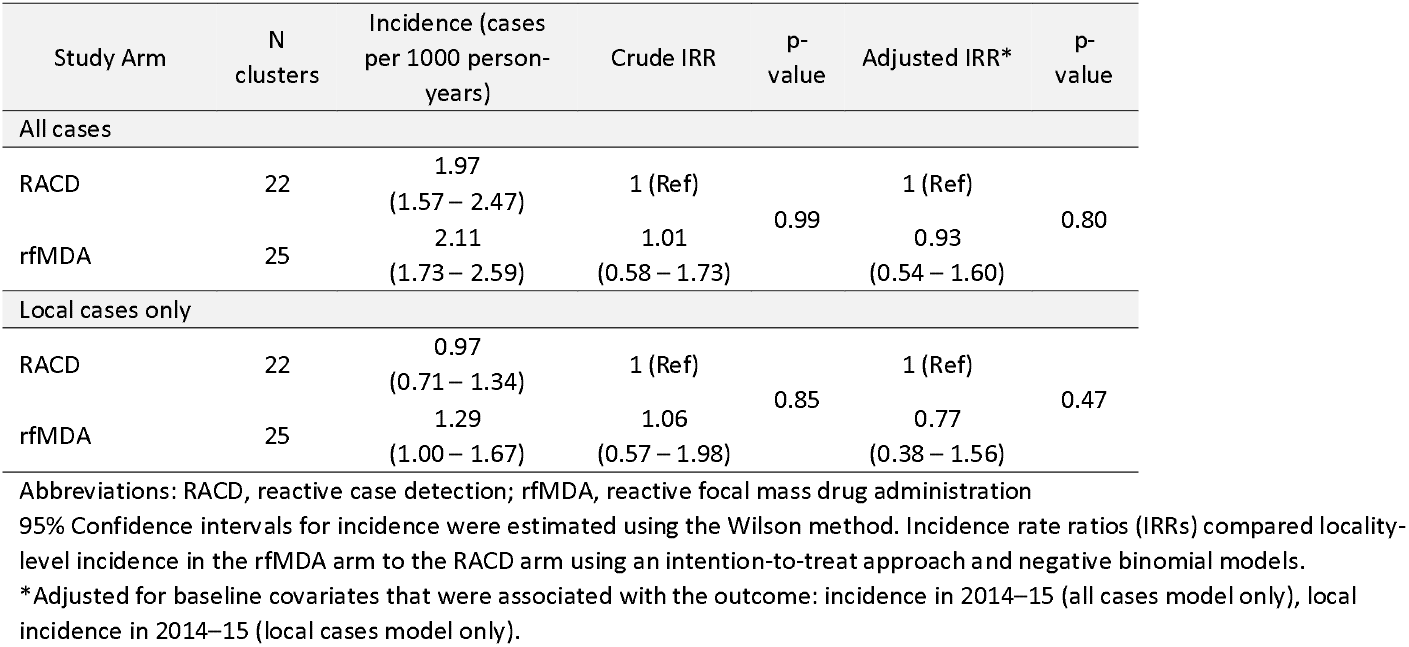
Adjusted incidence rate ratios (IRRs) in 2015–2017 comparing clusters assigned to RACD versus rfMDA

| Study Arm | N clusters | Incidence (cases per 1000 person-years) | Crude IRR | p-value | Adjusted IRR* | p-value |
| --- | --- | --- | --- | --- | --- | --- |
| All cases |  |  |  |  |  |  |
| RACD | 22 | 1.97<br>(1.57 – 2.47) | 1 (Ref) | 0.99 | 1 (Ref) | 0.80 |
| rfMDA | 25 | 2.11<br>(1.73 – 2.59) | 1.01<br>(0.58 – 1.73) |  | 0.93<br>(0.54 – 1.60) |  |
| Local cases only |  |  |  |  |  |  |
| RACD | 22 | 0.97<br>(0.71 – 1.34) | 1 (Ref) | 0.85 | 1 (Ref) | 0.47 |
| rfMDA | 25 | 1.29<br>(1.00 – 1.67) | 1.06<br>(0.57 – 1.98) |  | 0.77<br>(0.38 – 1.56) |  |

Abbreviations: RACD, reactive case detection; rfMDA, reactive focal mass drug administration 95% Confidence intervals for incidence were estimated using the Wilson method. Incidence rate ratios (IRRs) compared locality-level incidence in the rfMDA arm to the RACD arm using an intention-to-treat approach and negative binomial models. \*Adjusted for baseline covariates that were associated with the outcome: incidence in 2014–15 (all cases model only), local incidence in 2014–15 (local cases model only).

**Figure 3.**
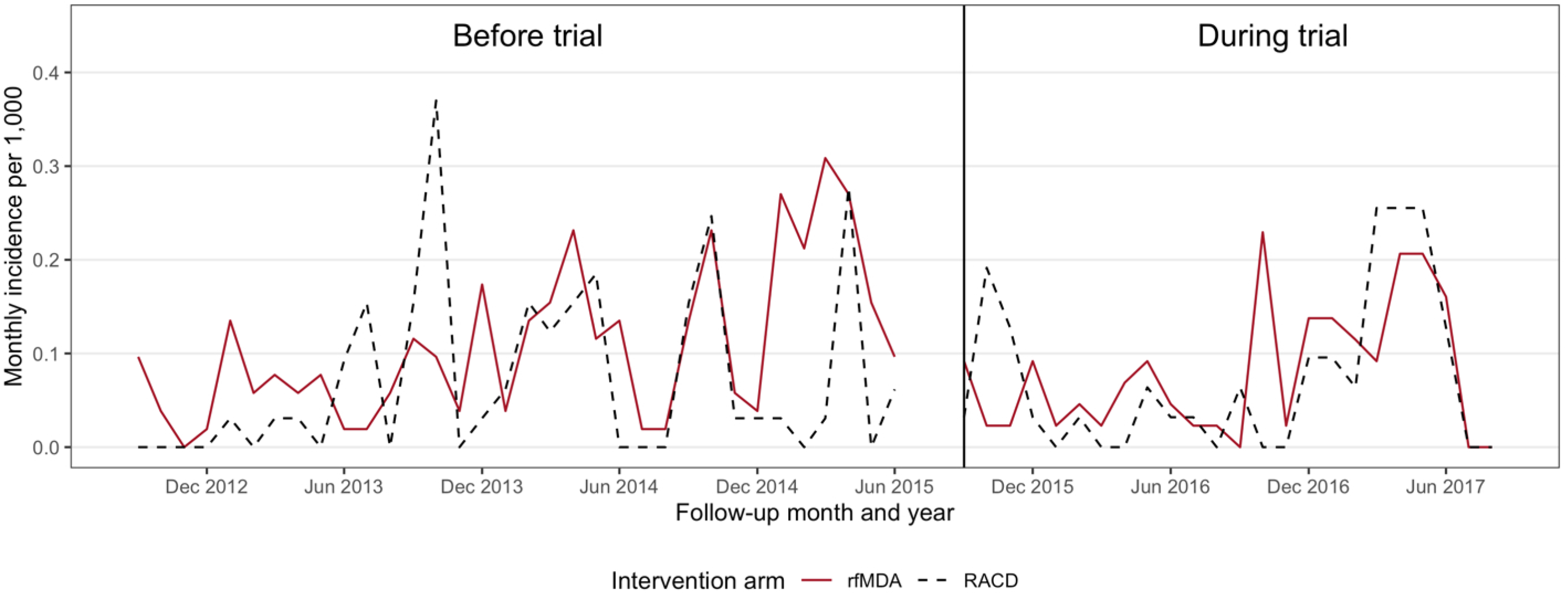
Monthly incidence in each study arm prior to and during the intervention period. RACD, reactive case detection; rfMDA, reactive focal mass drug administration

Cumulative malaria-free survival for all cases was similar between arms (Figure 4a). Restricting to local cases only, cumulative survival was higher in the rfMDA arm until approximately 9 months after study initiation, and subsequently it was higher in the RACD arm throughout the second high transmission season (13–18 months after study initiation) (Figure 4b). The Schoenfeld residual test indicated that survival was proportional between arms for all cases (rho = −0.04, p-value = 0.308) but not for local cases (rho = 0.12, p-value = 0.032).

**Figure 4.**
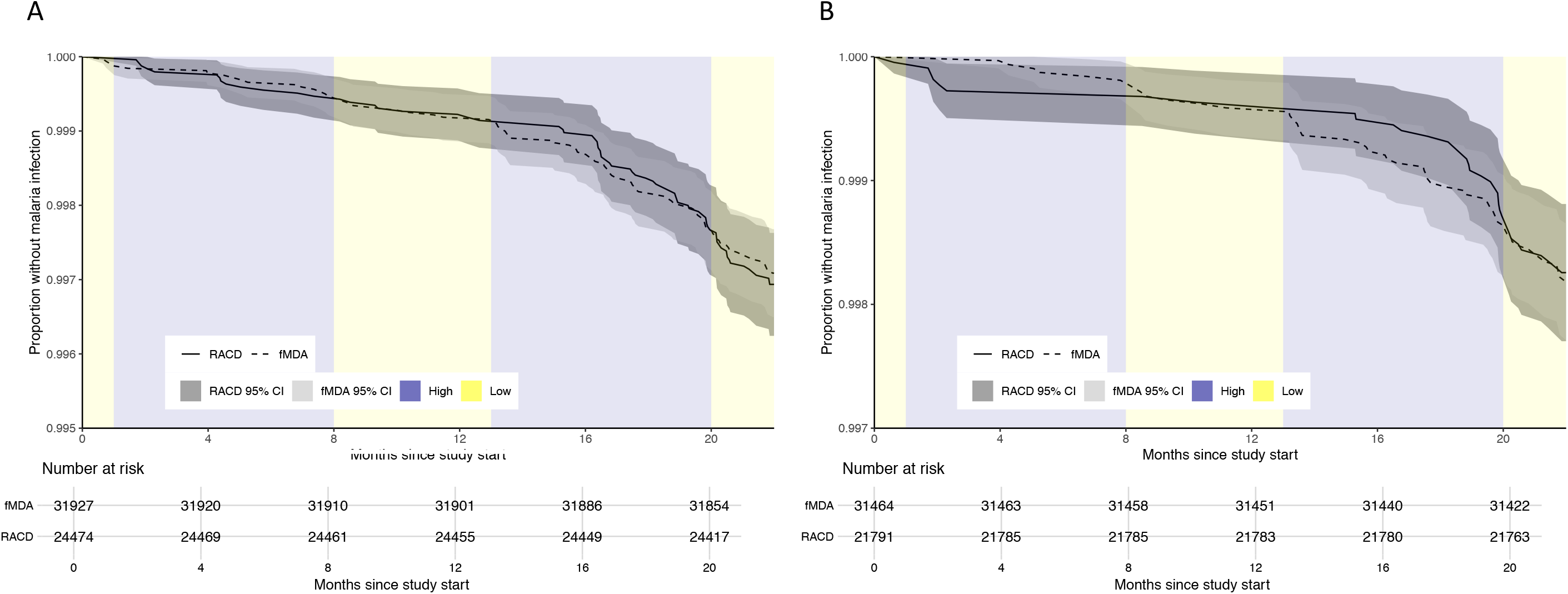
Malaria-free survival curves for the outcomes of a) all incident malaria cases, and b) local incident malaria cases. RACD, reactive case detection; rfMDA, reactive focal mass drug administration. High transmission seasons occurred from follow-up months 1-5 and 13-18.

In the synthetic control analysis accounting for pre-trial differences in incidence between arms, there was no difference in incidence of all malaria cases between the rfMDA arm and the synthetic RACD arm (Appendix 5 and 6). When comparing incidence in each cluster to incidence in contiguous neighbouring clusters, cluster-level incidence was not associated with incidence in contiguous neighbouring clusters, suggesting that the risk of contamination in this trial was minimal (Appendix 7).

Field staff conducted 1114 pill counts and recorded complete adherence to the 3-day DP regimen in 1099 (98.7%) individuals. Adverse events were experienced by 68 individuals in the rfMDA arm (49 in year 1, 19 in year 2). Based on the WHO severity scale, 54 (80%) events were mild and 14 (20.6%) were as moderate. The most common complaints were headache, nausea/vomiting, and abdominal pain. Of five individuals with adverse events who did not complete the course of DP, all recovered. One had difficulty breathing and chest tightness that could be consistent with DP-associated arrythmia but the accompanying diarrohea is less consistent (Appendix 8 and 9). During the study period, there was only one recorded malaria death in the study area. The infection was locally acquired and the patient lived in an rfMDA cluster, though rfMDA had not previously been conducted in the target area. No AEs were reported in the RACD arm.

## Discussion

In this pragmatic, cluster-randomised trial conducted in a very low transmission malaria elimination setting, rfMDA clusters had lower locally acquired malaria incidence during the whole study period compared to RACD clusters, particularly during the second high transmission season of the study period, but overall, evidence was weak. Intervention coverage was lower than expected, and malaria occurred in fewer clusters than planned in the sample size calculation. Adherence to presumptive treatment with DP was high, and as reported elsewhere, acceptability was high.(18) Importantly, there were no serious adverse events (SAEs).

Progress towards the 2030 elimination goal in southern Africa has slowed despite coordinated regional efforts and delivery of standard interventions, including pre-season indoor residual spraying, symptomatic case management, and RACD.(11) While RACD aims in part to address asymptomatic reservoirs of transmission, rapid diagnostic tests used in low transmission settings have poor sensitivity and miss many low-density infections.(24) While blanket MDA would reach all asymptomatic infections, it is logistically difficult to implement at scale, inefficient in populations with few, highly clustered infections, and it may not be safe or acceptable.(7)

A few trials have evaluated focal MDA delivered to hotspots at the village or sub-village level in low transmission settings and results are mixed.(25-28) In Zanzibar, which most resembles our site due to high coverage of standard interventions, very low transmission intensity, and high rates of importation, negative findings of focal MDA effectiveness were hypothesized to be related to suboptimal timing and the number of MDA rounds, and re-introduction of malaria through importation.(27) The reactive approach employed in our trial sought to address these issues by targeting the focal MDA to a time and place when transmission risk was highest (e.g. where there were recent imported or local cases).

This trial is one of three that evaluated rfMDA. Results from a low transmission setting in Zambia trial are forthcoming.(25) A trial in a low transmission setting in Namibia evaluated rfMDA alone and in combination with reactive vector control in comparison to RACD.(13) Compared to RACD, rfMDA reduced local malaria incidence by 48%, and rfMDA with additional reactive vector control reduced incidence by 74%. There are several key differences between the Namibia trial and this trial. First, the Namibia trial had a higher baseline annual malaria incidence (30 per 1000 compared to 3 per 1000 in this trial) and a lower proportion of imported malaria (3% compared to 40% in this trial), both of which may facilitate higher impact of focal MDA.(26) Second, the Namibia trial was largely implemented by a research team, while the Eswatini trial was pragmatic and largely implemented by the local malaria control programme.(16) Coverage in the Namibia trial was also higher compared to this trial (study area index and target population coverage were >84% and >85%, compared to 78.5% and 78.7% in this trial).

This trial faced several challenges unique to very low incidence settings including strong spatiotemporal clustering and imported malaria.(22) The number of clusters with at least one index case during follow-up was lower than expected (we expected 63 but observed 47). Thus, the trial was not powered to detect the hypothesized incidence reduction of ≥50%, nor smaller reductions, with precision. Second, though the study was cluster-randomised, baseline malaria incidence and the percentage of imported cases was higher in the rfMDA arm than the RACD arm. The lower than expected number of clusters per arm likely contributed to these imbalances. Though analyses adjusted for these factors, it remains possible that unmeasured factors affecting malaria transmission differed between arms. To further address baseline imbalance, we conducted a synthetic control analysis, which produced similar results to the primary analysis. However, the synthetic control analysis did not completely account for pre-intervention differences in incidence between arms, likely because few clusters (<25 per arm) had incident cases during follow-up, and cluster-level incidence varied. When outcomes are rare and clustered, trials require very large cluster numbers to have sufficient statistical power and baseline balance.(27, 28)

Implementation factors may have influenced effect estimates. First, total coverage was lower than the trial’s goal of 80%, and imbalanced across arms (60% for RACD and 68.8% for rfMDA). Limitations related to staffing and transport compromised index case level coverage and participants not being present compromised target population coverage. Of note, 12% of the rfMDA target population was ineligible to receive DP, with the most common reason being potential medication interaction with ARVs. Although saquinavir, the only ARV contraindicated for use with DP, is not available in Eswatini, nurses expressed concern that adverse events could be interpreted by the participant as due to ARV, and thus compromise ARV adherence. Where ARV use is common, such as Eswatini which has the highest worldwide incidence of human immunodeficiency virus (HIV),(29) better strategies to address safety concerns regarding drug-drug interaction will be needed. The use of less stringent exclusion criteria (e.g. inclusion of pregnant women, young children, individuals with certain morbidities) as has been safely practiced by others(30) could also improve coverage. However, coverage was not associated with incidence, suggesting that differences in coverage between arms were unlikely to affect study findings. Second, intervention response time was substantially higher for two rfMDA clusters compared to the RACD arm. It is possible that there was greater malaria transmission between index case detection and intervention delivery in the rfMDA arm than the RACD arm. Third, study clusters were not separated by geographic buffer zones to minimize contamination, which can occur due to vectors or human movement. However, cluster-level incidence was not correlated between contiguous clusters, suggesting that the chance of contamination in this trial was low.

Importantly, our study is the first to show the safety of rfMDA using DP. The Namibia trial used AL and in comparison, DP may be preferable for MDA due to ease of use and longer protective period (once versus twice daily, and 4–6 weeks versus a few days, due to the half-life). Rarely, DP-associated QT-interval prolongation may lead to sudden death (1/∼200 000), and in very low-endemic settings the risk-to-benefit ratio may not favor DP.(15) Here, one participant had symptoms that could be consistent with arrythmia, and treatment was stopped. Pharmacovigilance provided by nurses through follow-up visits and their on-call availability likely helped to prevent SAEs.

## Conclusions

This study is the first trial to compare rfMDA and RACD in a very low malaria-endemic setting. As interventions were embedded within an existing national malaria programme, it provides such evidence in realistic implementation conditions. We found that rfMDA was safe. Although rfMDA clusters had lower cumulative incidence during the study period, we were unable to confirm effectiveness of rfMDA compared to RACD, potentially due to insufficient power. For rfMDA to be more effective than RACD, improved coverage and/or the addition of complementary interventions, such as IRS, may need to be delivered in tandem.(13) To improve statistical power to detect impact of interventions in very low-endemic settings, future trials may require multi-site designs, larger sample sizes, or alternatively, smaller units of randomisation (e.g. a neighborhood), or adaptive designs that adjust features such as the sample size and allocation ratio.(31, 32) Such evidence will be critical to guide countries in their quest to move from very low to no transmission.

## Data Availability

The data that support the findings of this study are available from Eswatini Ministry of Health but restrictions apply to the availability of these data, which were used under license for the current study, and so are not publicly available. Data are however available from the authors upon reasonable request and with permission of Eswatini Ministry of Health. 

## List of abbreviations

aITT: adjusted intention to treat
ARV: antiretrovirals
DP: dihydroartemisinin-piperaquine
HIV: human immunodeficiency virus
ITT: intention to treat
IRS: indoor residual spraying
LAMP: loop mediated isothermal amplification
MDA: mass drug administration
NMP: National Malaria Programme
PCR: polymerase chain reaction
RACD: reactive case detection
rfMDA: reactive focal mass drug administration
RDT: rapid diagnostic test
SAE: serious adverse events
WHO: World Health Organization

## Declarations

### Ethics approval and consent to participate

Ethics approval was given by Eswatini Ministry of Health (MH/599C) and by University of California San Francisco Human Research Protection Program & IRB (Formerly Committee on Human Research) (14-15226). Written informed consent was obtained from individual participants. For children less than 18 years, written informed consent from a parent or guardian was required, as was written assent for children 12–17 years.

### Trial registration

ClinicalTrials.gov, NCT02315690 (registration date: December 8, 2014)

### Consent for publication

Not applicable

### Patient and Public Involvement

As assessed formally (through Knowledges Attitudes and Practices surveys) and informally (during malaria programme activities including reactive case detection), the public’s concerns about malaria and their eagerness for the country to achieve its goal of malaria elimination informed the research question and study design. As incident malaria cases were the trigger for recruitment (targeting household members and neighbors of index cases) and the primary outcome, these aspects of the study relied on patients seeking care when ill and receiving malaria testing. To elicit ongoing feedback regarding the conduct and burden of the study intervention, patients and the public were engaged in an ongoing basis through focus group discussions, the results of which are published elsewhere.(18)

### Authors’ contributions

MSH, SK, and RG conceptualised and designed the study. NN, ND, and KB contributed to study design. NM led the trial coordination. KB, BD, DH, LMP, and CM additionally supported trial coordination. ND led the field implementation. SV led the data collection. MK and GT oversaw clinical and safety aspects of the trial. KB oversaw data collection and analysis of acceptability assessment. NN led the laboratory activities with oversight from DH, BG, and GM. SV and BW led data management and supported data analyses. MSKD, JBC, and MSH led the data analysis. RG and DP advised on the data analyses. JBC and MSH wrote the manuscript. NN and SK provided oversight of local implementation. MSH provided overall oversight of the study. All authors read and approved the final manuscript.

## Acknowledgements

The authors would like to thank the residents of Eswatini who supported the study through their participation and inputs. We thank the field and laboratory staff. We thank Alemayehu for collecting ecological data. We thank Adam Soble, Manik Saini, Charlotte Lejeune, and Thomas How at CHAI for their support in administration and local coordination. We thank Justin Cohen, Arnaud LeMenach, Hugh Sturrock, Joelle Nadle, Immo Kleinschmidt, and Robert Haley for their inputs on trial design. We thank the Ministry of Health, Eswatini Pharmacovigilance committee, and the Eswatini Malaria Elimination Advisory Group for their support and oversight.

