## Supplemental for "Effectiveness and safety of reactive focal mass drug administration (rfMDA) using dihydroartemisinin-piperaquine to reduce malaria transmission in very low-endemic setting of Eswatini: a pragmatic cluster randomised controlled trial"

### **Appendix 1. Inclusion and exclusion criteria**

|  | **Inclusion criteria** | **Exclusion criteria** |
| --- | --- | --- |
| Index case (as trigger for RACD or rfMDA in their household members and neighbours) | - Laboratory-confirmed malaria case (either locally transmitted or imported) detected at and reported from a health facility, and - Resided in a study locality. | - Malaria infection identified through RACD or other active case detection. |
| RACD intervention | - Non-index case, and - Resided or spent at least one night in the Target Area in the past 5 weeks. | - Target Area overlaps with prior Target Area that received the RACD intervention within the past 5 weeks. |
| rfMDA intervention | - Non-index case, and - Resided or spent at least one night in the Target Area in the past 5 weeks. | - Target Area overlaps with prior Target Area that received the TPE intervention within the past 8 weeks, and - For dihydroartemisinin-piperaquine (DP) specifically (though still eligible for interview): - Temperature ≥ 38·0⁰C, report of fever in the past 48 hours, or other illness (will be referred to the nearest health facility for further evaluation) - Reported pregnancy or breastfeeding in women who have had menarche but no menses in the past 4 weeks (assessed in women and girls ≥10 years of age) - Children <9 months of age or <7 kg - Known allergy or history of adverse reaction to DP - Already taken 2 courses of DP in the past year or taken 1 course within the past 2 months - Moderate or severe renal or hepatic insufficiency - Currently with severe malaria - Family history of sudden death or of congenital prolongation of the QTc interval. - Known congenital prolongation of the QTc-interval or any clinical condition known to prolong the QTc interval. - History of symptomatic cardiac arrhythmias or with clinically relevant bradycardia. Any predisposing cardiac conditions for arrhythmia such as severe hypertension, left ventricular hypertrophy (including hypertrophic cardiomyopathy) or congestive cardiac failure accompanied by reduced left ventricle ejection fraction. - Electrolyte disturbances, particularly hypokalaemia, hypocalcaemia or hypomagnesaemia (including vomiting in child) - Recent treatment with medicinal products known to prolong the QTc interval that may still be circulating at the time that DP is commenced (e.g. mefloquine, halofantrine, lumefantrine, chloroquine, quinine and other antimalarial agents) |

Abbreviations: RACD, reactive case detection; rfMDA, reactive focal mass drug administration The Target Area is defined for rfMDA localities as all individuals residing within 200m of an index case that is detected in passive surveillance and resides in a rfMDA study locality, with individuals residing immediately beyond 200m included if a minimum of 30 individuals are not enrolled within 200m. The Target Area is defined for RACD localities as all individuals residing within 500m of an index case that is detected in passive surveillance and resides in a RACD locality.

### **Appendix 2. Methods for synthetic control analysis**

Because cluster-level monthly incidence was not balanced between study arms at baseline, we used a synthetic control analysis to create a synthetic RACD arm that better approximated the counterfactual level of cluster-level incidence in the RACD arm had the study arms been balanced. The objective was to estimate the difference in incidence between arms had the trial been balanced at baseline.

The synthetic control arm consists of a weighted combination of actual RACD cluster data. We estimated monthly incidence in the synthetic RACD arm as a weighted average of observed monthly incidence in the RACD arm using weights that minimized the squared error between observed pre-trial monthly incidence in the rfMDA and RACD arm using nonlinear optimization. We constrained weights such that they were >0 and summed to 1. Clusters received larger weights if they better represented pre-intervention monthly incidence in the rfMDA arm. RACD localities with larger pre-trial monthly incidence tended to receive smaller weights, and other localities received weights that were smaller or equal to zero (Appendix 2 Figure 1). Clusters that contributed to the synthetic RACD arm had less variable pre-intervention monthly incidence than those that were excluded from the synthetic RACD arm (because their weights equaled 0) (Appendix 2 Figure 2).

Compared to the observed RACD pre-intervention incidence, the synthetic RACD pre-intervention incidence more closely mimicked the rfMDA pre-intervention incidence (Appendix 6). However, the synthetic RACD arm did not completely resemble pre-intervention monthly incidence in the rfMDA arm. This is likely because the study included a relatively small number of clusters per arm, and incidence varied substantially between arms, making it more difficult for the algorithm to identify RACD clusters with similar pre-intervention incidence to rfMDA clusters. We then compared the difference in observed incidence during trial follow-up in the rfMDA arm to the expected difference in the synthetic RACD arm.

To compare the two arms, we estimated the cumulative incidence prior to and during the trial in the synthetic RACD arm standardizing by person-months in the rfMDA arm. Because the pre-intervention differences in incidence remained between the synthetic RACD arm and the rfMDA arm, we used a difference-in-difference approach, which was calculated as follows to compare rfMDA to RACD: [(rfMDA incidence during the intervention − rfMDA incidence pre-intervention) − (RACD incidence during the intervention − RACD incidence pre-intervention)]. We calculated the analogous difference-in-difference to compare the rfMDA to the synthetic RACD arm.

**Appendix 2 Figure 1. Monthly incidence prior to and during the study intervention in each reactive case detection (RACD) locality that received a non-zero weight in the synthetic control analysis**

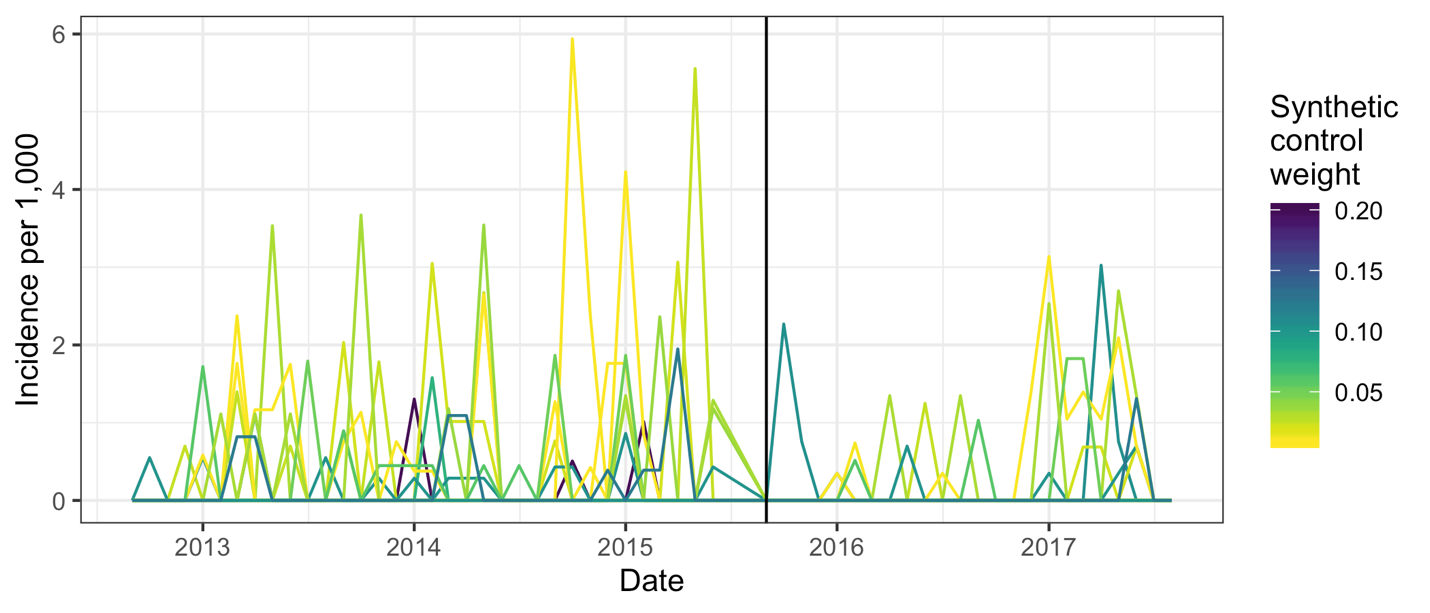

The synthetic control method assigned weights > 0 to 16 out of 22 localities in the RACD arm.

**Appendix 2 Figure 2. Monthly incidence prior to and during the study intervention in each study locality and in the synthetic reactive case detection (RACD) group**

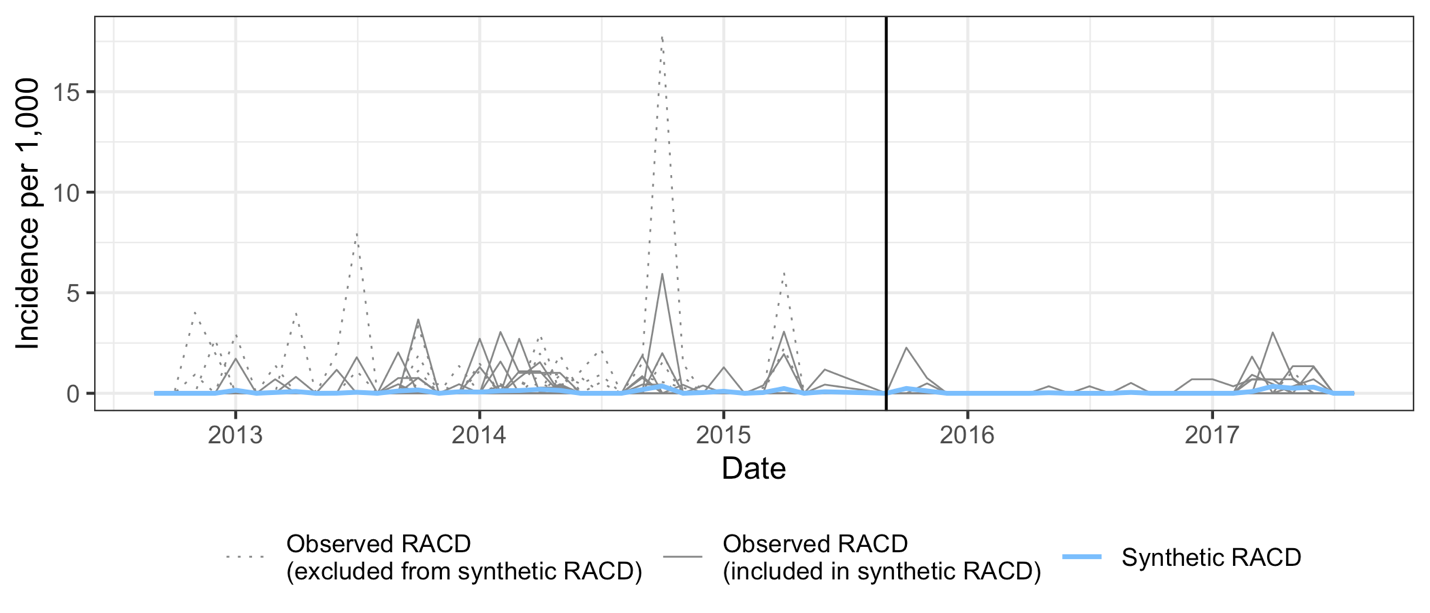

### **Appendix 3. Characteristics of index cases and target population, mean % across clusters (95% CI) during two-year follow-up**

| Locality or cluster-level characteristic | Overall  n=47* | RACD  n=22** | rfMDA  n=25*** |
| --- | --- | --- | --- |
| INDEX CASES |  |  |  |
| Male | 48·5 (38·4 – 58·5) | 48·5 (32·4 – 64·7) | 48·5 (34·8 – 62·1) |
| Age (years) |  |  |  |
| <15 | 15·3 (8·9 – 21·6) | 15·8 (7·1 – 24·4) | 14·8 (5·0 – 24·6) |
| 15–40 | 60·2 (50·4 – 70·0) | 59·7 (43·4 – 76·0) | 60·6 (48·0 – 73·3) |
| >40 | 22·3 (13·4 – 31·3) | 24·4 (8·6 – 40·1) | 20·5 (9·8 – 31·3) |
| International travel in past 8 weeks | 35·7 (25·6 – 45·9) | 37·6 (20·5 – 54·7) | 34·1 (21·0 – 47·2) |
| Occupation  Agricultural  Manual labor  Manufacturing  Office  Small market sales/trade  Unemployed/Retiree  Student  Other  Child, non-student | 17·7 (8·7 – 26·6)  7·8 (2·3 – 13·2)  2·4 (-0·22 – 5·0)  3·0 (0·22 – 5·8)  5·4 (1·8 – 9·0)  35·2 (24·1 – 46·4)  21·7 (13·0 – 30·4)  1·4 (0·19 – 2·6)  3·2 (1·1 – 5·2) | 20.3 (6.4 – 34.2)  5.7 (-0.24 – 11.7)  3.1 (-1.7 – 7.9)  4.3 (-1.1 – 9.7)  5.5 (-0.22 – 11.3)  42.3 (23.7 – 60.9)  13.9 (5.4 – 22.5)  1.0 (-0.86 – 2.9)  3.6 (0.28 – 7.0) | 15.3 (2.9 – 27.8)  9.6 (0.55 – 18.6)  1.8 (-1.1 – 4.6)  1.9 (-0.81 – 4.6)  5.3 (0.45 – 10.2)  29.0 (15.0 – 43.0)  28.6 (14.0 – 43.1)  1.8 (0.05 – 3.5)  2.7 (0.01 – 5.4) |
| Home sprayed in the past year  Yes  No | 17.7 (10.1 – 25.4)  71.8 (62.0 – 81.5) | 5.3 (-0.68 – 11.3)  82.2 (68.2 – 96.1) | 28.6 (16.4 – 40.8)  62.6 (49.2 – 76.0) |
| Own bednet  Yes  No | 22.9 (13.9 – 31.9)  74.9 (65.3 – 84.4) | 20.0 (8.1 – 31.8)  79.9 (68.0 – 91.8) | 25.6 (11.5 – 39.6)  70.4 (55.3 – 85.6) |
| TARGET POPULATION |  |  |  |
| Male | 46.1 (41.3 – 50.9) | 40.4 (34.3 – 46.5) | 51.3 (44.3 – 58.4) |
| Age (years) |  |  |  |
| <15 | 42.2 (37.3 – 47.1) | 42.2 (34.5 – 49.9) | 42.2 (35.2 – 49.1) |
| 15–40 | 40.1 (34.8 – 45.3) | 41.5 (32.8 – 50.1) | 38.8 (31.8 – 45.8) |
| >40 | 17.7 (14.8 – 20.7) | 16.3 (12.3 – 20.3) | 19.0 (14.4 – 23.6) |
| International travel | 2.8 (0.36 – 5.1) | 2.3 (0.19 – 4.4) | 3.2 (-1.2 – 7.5) |
| Occupation  Agricultural  Manual labor  Manufacturing  Office  Small market sales/trade  Unemployed/Retiree  Student  Other  Child, non-student | 15.8 (7.3 – 24.4)  2.8 (1.6 – 3.9)  0.80 (0.32 – 1.3)  1.5 (0.75 – 2.3)  2.3 (1.4 – 3.3)  28.6 (23.7 – 33.4)  31.9 (26.4 – 37.4)  2.7 (-0.91 – 6.3)  13.5 (10.7 – 16.4) | 9.5 (-1.5 – 20.5)  3.2 (0.94 – 5.4)  0.10 (-0.02 – 0.23)  1.4 (0.36 – 2.4)  2.5 (0.86 – 4.2)  32.8 (25.9 – 39.8)  29.6 (21.8 – 37.5)  4.4 (-3.4 – 12.2)  16.3 (11.4 – 21.2) | 21.6 (8.2 – 34.9)  2.4 (1.2 – 3.6)  1.4 (0.58 – 2.3)  1.6 (0.42 – 2.9)  2.2 (0.93 – 3.4)  24.7 (17.8 – 31.6)  34.0 (25.8 – 42.1)  1.2 (-0.08 – 2.4)  11.0 (7.8 – 14.1) |
| Household sprayed in past year (individual level) |  |  |  |
| Yes | 26.5 (16.2 – 36.8) | 19.5 (5.4 – 33.6) | 32.9 (17.3 – 48.5) |
| No | 69.0 (58.0 – 79.9) | 77.0 (62.9 – 91.2) | 61.7 (44.7 – 78.7) |
| Don’t know | 4.4 (1.6 – 7.2) | 3.3 (0.50 – 6.2) | 5.4 (0.55 – 10.2) |
| Household sprayed in past year (household level) |  |  |  |
| Yes | 25.2 (15.4 – 35.0) | 21.6 (7.8 – 35.4) | 28.5 (13.6 – 43.3) |
| No | 69.2 (58.5 – 79.8) | 72.9 (58.5 – 87.3) | 65.7 (49.1 – 82.5) |
| Don’t know | 5.6 (2.4 – 8.8) | 5.5 (1.8 – 9.3) | 5.7 (0.34 – 11.1) |

Abbreviations: RACD, reactive case detection; rfMDA, reactive focal mass drug administration.

Data are mean proportion (95% CI).

* n=40 for target population. N for target population is smaller than for index population because some index cases did not get an intervention response (index case coverage was not 100%).

** n=19 for target population. N for target population is smaller than for index population because some index cases did not get an intervention response (index case coverage was not 100%).

*** n=21 for target population. N for target population is smaller than for index population because some index cases did not get an intervention response (index case coverage was not 100%).

### **Appendix 4. Locality-level incidence in the year prior to the trial (A) and during the trial (B) by intervention arm**

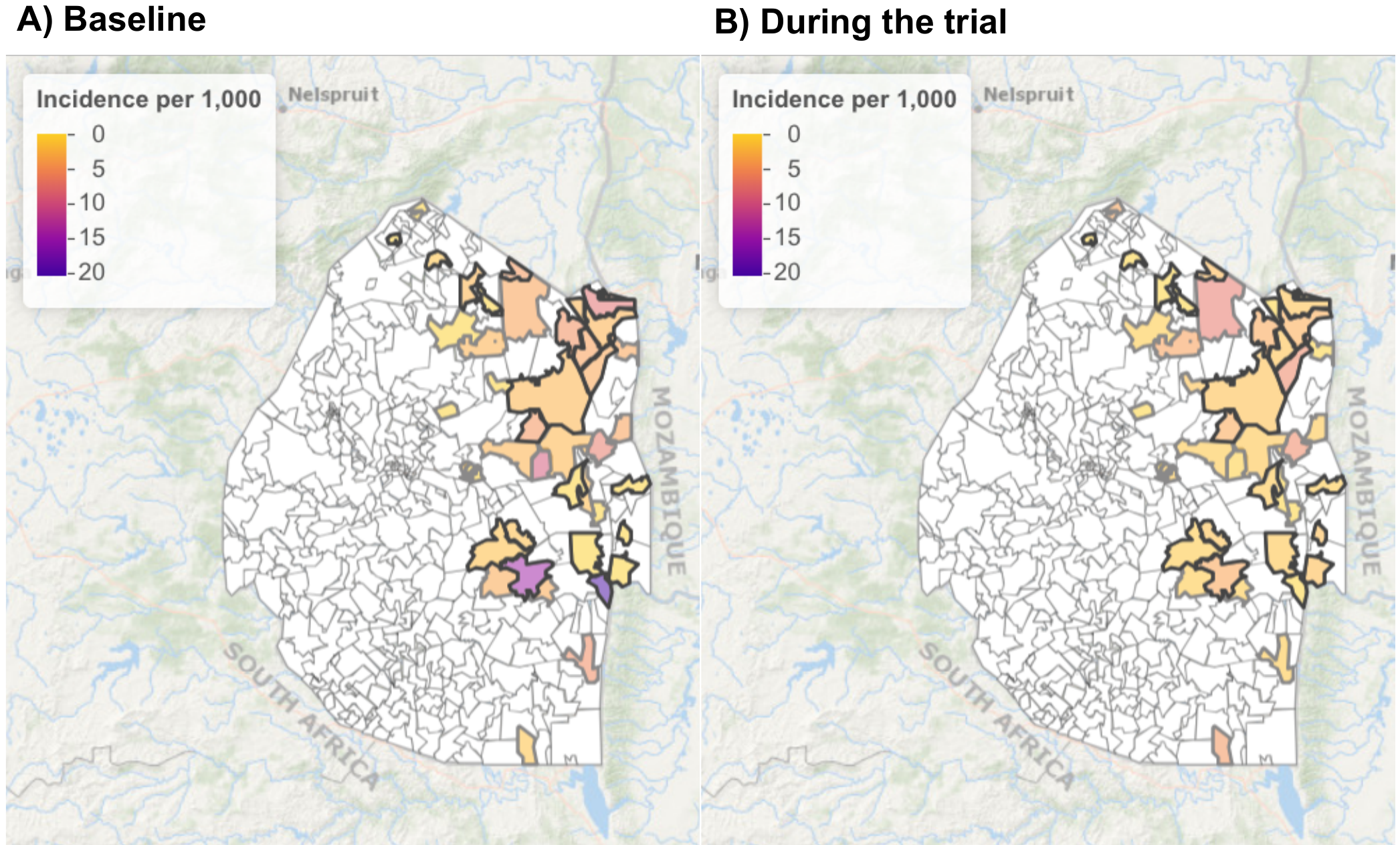

Map of the study area with malaria incidence per 1 000 by locality. rfMDA localities have black outlines and RACD localities have grey outlines. Localities not included in the trial are shown in white.

### **Appendix 5. Monthly incidence prior to the intervention in each study arm and in the synthetic control group**

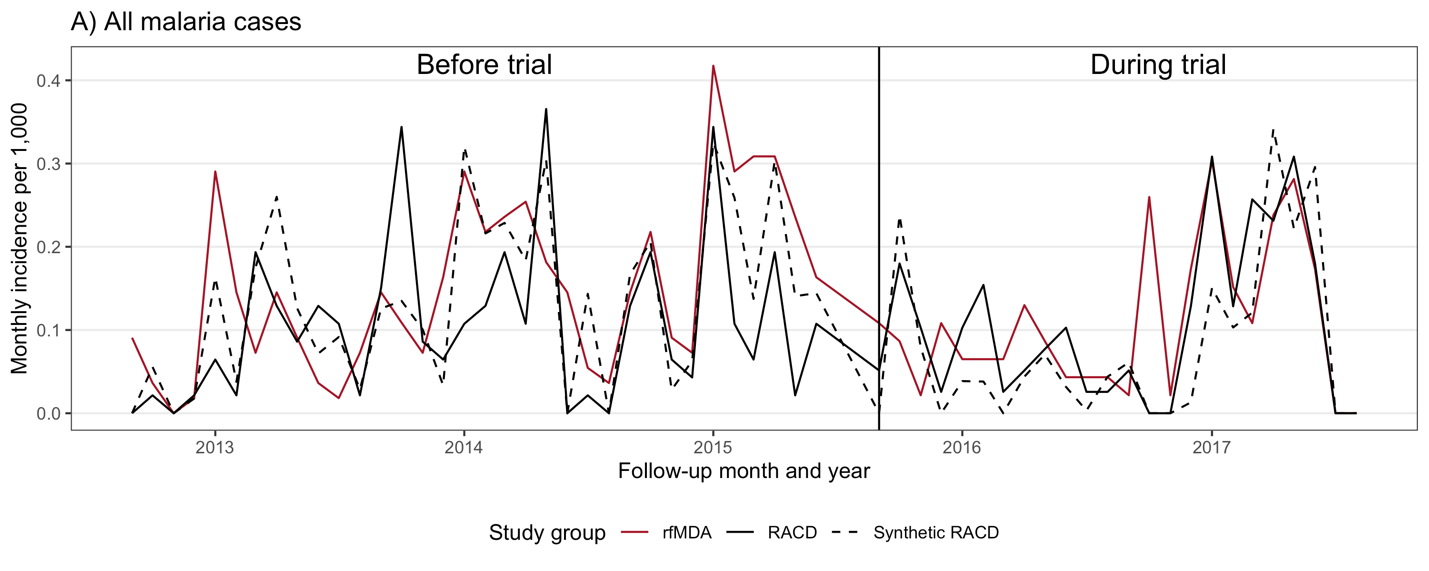

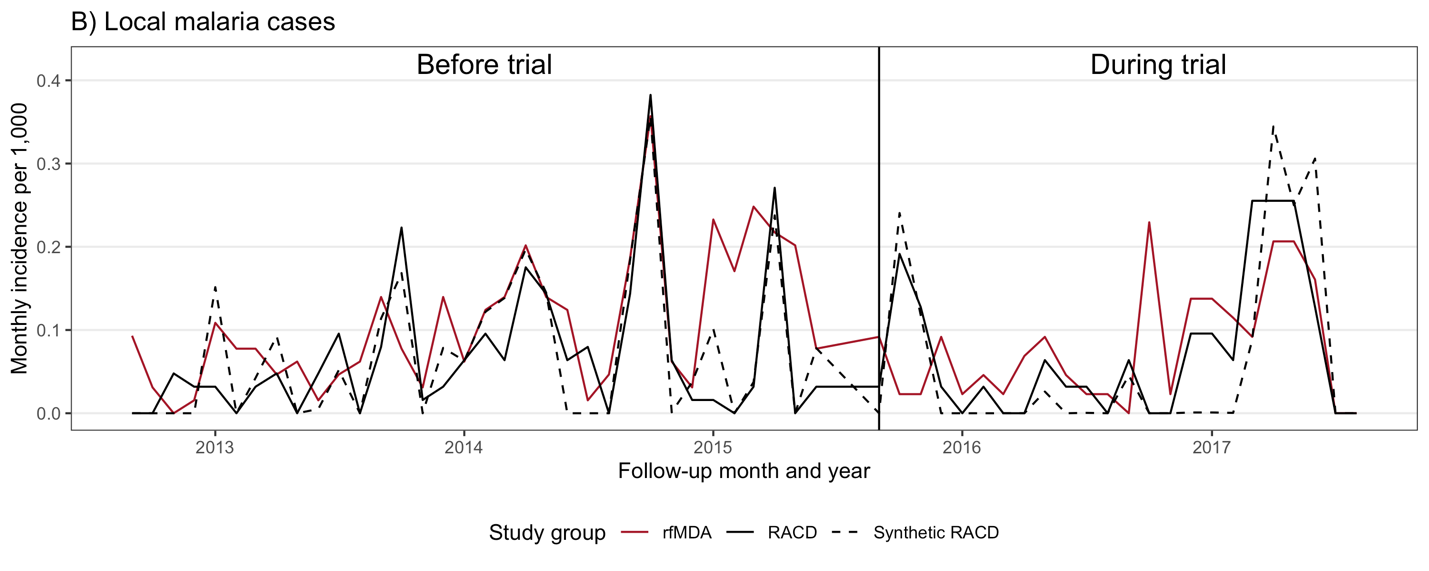

Abbreviations: RACD, reactive case detection; rfMDA, reactive focal mass drug administration.

### **Appendix 6. Incidence differences in the primary analysis and the synthetic control analysis**

| **Study group** | **Pre-intervention** | | | | **During Intervention** | | | | **Difference-in-difference** |
| --- | --- | --- | --- | --- | --- | --- | --- | --- | --- |
|  | **Cases** | **Person-months** | **Incidence per 1 000** | **Incidence difference** | **Cases** | **Person-months** | **Incidence per 1 000** | **Incidence difference** |  |
| **All malaria cases** | | | | | | | | | |
| rfMDA | 130 | 23,316 | 5.6 | -- | 84 | 32,179 | 2.6 | -- | -- |
| RACD | 88 | 23,316 | 3.8 | 1.8 | 81 | 32,179 | 2.5 | 0.1 | -1.7 |
| Synthetic RACD | 113 | 23,316 | 4.8 | 0.7 | 61 | 32,179 | 1.9 | 0.7 | 0.0 |
| **Local malaria cases** | | | | | | | | | |
| rfMDA | 93 | 23,316 | 4.0 | -- | 61 | 32,179 | 1.9 | -- | -- |
| RACD | 58 | 23,316 | 2.5 | 1.5 | 56 | 32,179 | 1.8 | 0.1 | -1.4 |
| Synthetic RACD | 60 | 23,316 | 2.6 | 1.4 | 46 | 32,179 | 1.4 | 0.5 | -0.9 |

Abbreviations: RACD, reactive case detection; rfMDA, reactive focal mass drug administration.

Incidence differences were calculated as the incidence in the rfMDA arm minus the incidence in the RACD arm or the synthetic RACD arm. Difference-in-differences for the RACD arm were calculated as the [(rfMDA incidence during the intervention − rfMDA incidence pre-intervention) − (RACD incidence during the intervention − RACD incidence pre-intervention)], and analogously for the synthetic RACD arm.

### **Appendix 7. Locality level-incidence in contiguous localities by arm**

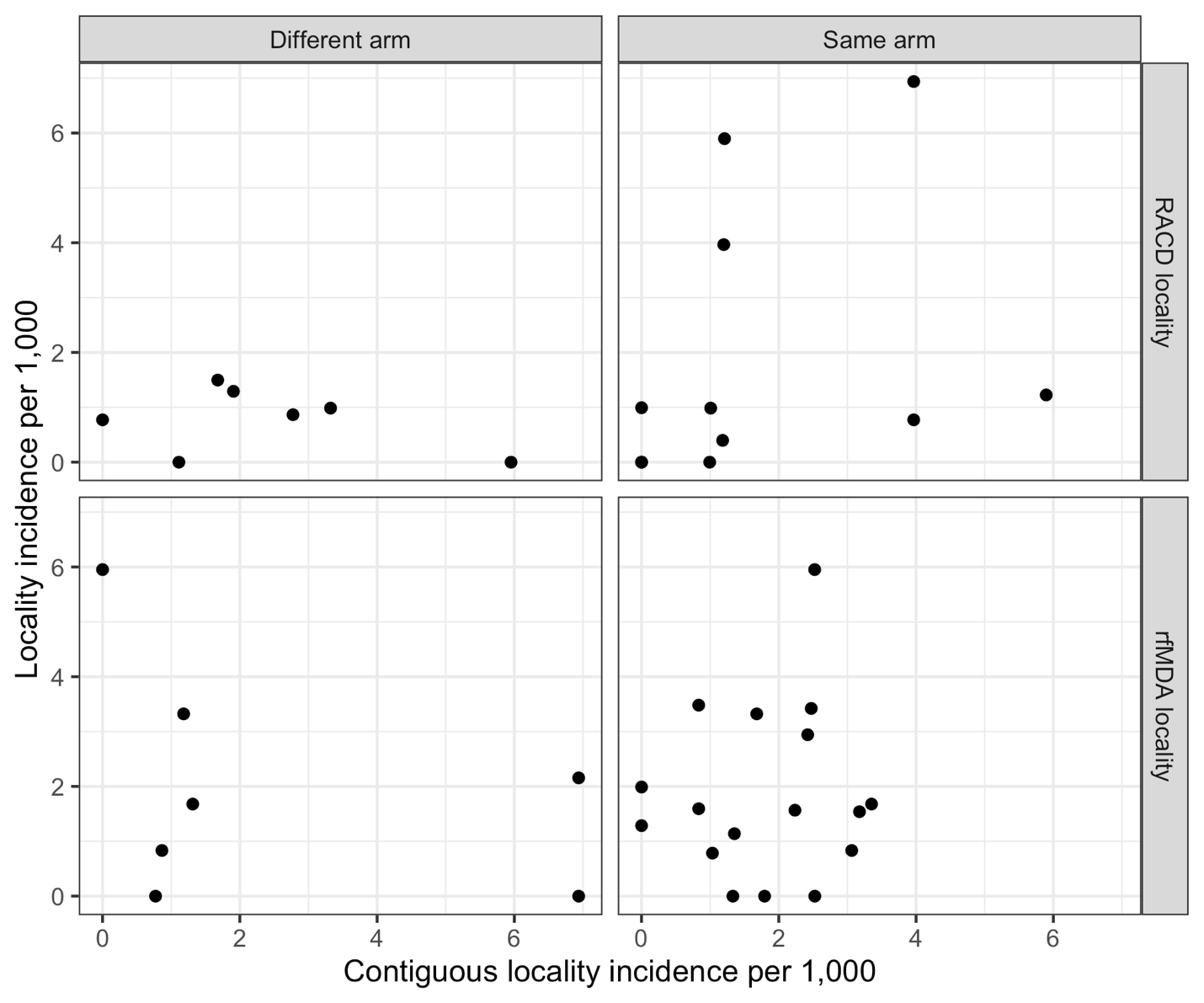

Abbreviations: RACD, reactive case detection; rfMDA, reactive focal mass drug administration.

### **Appendix 8. Symptomology of reported adverse events**

| Symptom | N=1,932*  n (%) |
| --- | --- |
| Headache | 37 (1.9) |
| Nausea and/or vomiting | 25 (1.3) |
| Abdominal pain | 17 (0.9) |
| Body weakness | 11 (0.6) |
| Fever | 7 (0.4) |
| Diarrhoea | 7 (0.4) |
| Rash | 5 (0.3) |
| Dizziness | 4 (0.2) |
| Chest pain and/or breathing difficulty | 3 (0.2) |
| Cough | 2 (0.1) |
| Red, itchy eyes | 2 (0.1) |

*number of individuals who received dihydroartemisinin-piperaquine (DP)

### **Appendix 9. Symptomology in participants with reported adverse events that did not complete the entire course of dihydroartemisinin-piperaquine (n=5)**

| **Individual No.** | **Age range (years)** | **Sex** | **Symptoms** | **Suspected relationship to study drug** | **Decision to stop therapy (self, nurse)** | **Details of therapy discontinuation** | **Confirmed recovery** | **Time to recovery** |
| --- | --- | --- | --- | --- | --- | --- | --- | --- |
| 1 | 15–40 | Female | Vomiting | Possible | Nurse | Discontinued after vomitting with first and second doses. | Yes | 1 day |
| 2 | 15–40 | Female | Nausea, fatigue | Probable | Self | Discontinued after first dose due to impact on activities of daily living, including ability to attend work. | Yes | 2 days |
| 3 | >40 | Male | Difficulty breathing | Probable | Self | Discontinued after second dose when symptoms started. | Yes | A few hours |
| 4 | 15–40 | Female | Hyperventilation, difficulty breathing, chest tightening, chest pains, nausea, vomiting, stomach ache, diarrhoea | Definite | Nurse | Discontinued after second dose when nurse became aware of symptoms and participant newly disclosed history of hypertension. No hospitalisation was required. | Yes | Not recorded |
| 5 | <15 | Male | Nausea and vomiting | Possible | Self | Discontinued after nausea and vomiting with second dose. | Yes | A few hours |
